# Long-term restricted mean survival time survival of anterior large vessel occlusion strokes treated with endovascular thrombectomy

**DOI:** 10.1101/2025.02.12.25322189

**Authors:** Daniel James Wellington, William Diprose, Douglas Campbell, Jae Beom Hong, Luke Boyle, Peter Alan Barber

## Abstract

**Introduction:** Endovascular thrombectomy (EVT) has revolutionized the treatment of patients with large vessel occlusion (LVO) stroke. Three-month functional independence ranges between 33 and 60%, and mortality between 12 and 25%. Few studies have presented outcomes beyond five years and their reported odds or hazard ratios are difficult to apply clinically. Median survival is also challenging to interpret correctly. Restricted mean survival time (RMST) is a clinically intuitive metric that gives survival times up to specified cutoff points. We aimed to determine RMST at one, two and five years after EVT for anterior circulation stroke patients and evaluate survival impact of various risk factors.

**Methods:** This cohort study examined survival of patients treated with EVT for anterior circulation LVO stroke from 2011-2024 from the New Zealand national stroke database. Unadjusted RMST was determined for all age brackets. Cox regression adjusted survival curves were used to determine RMST and survival probabilities at one, two and five years for significant covariates.

**Results:** There were 1457 anterior circulation LVO patients treated with EVT included for analysis (median follow-up 8.9 years [95% CI 7.57 – inf]). At five years, 415 (28%) patients had died. Patients aged 90 years or older had a RMST of 1.64 years, and a five-year survival probability of 33% compared with a 97% survival probability for those aged 15-39 years. Five-year RMST survival differences were; independent level of function versus dependent (0.77 years), low comorbidity versus high (0.71 years), mild-moderate NIHSS versus higher (0.49 years), smaller infarct cores versus large (0.48 years), and good reperfusion versus (0.67 years).

**Discussion:** RMST provided clinically useful survival estimates and subgroup survival differences after EVT, especially in those of advanced age. RMST is easy to communicate, captures survival nuances, and can provide absolute differences that are not possible when outcomes are measured using median survival alone.

## Introduction

Endovascular thrombectomy (EVT) has revolutionized the treatment of patients with large vessel occlusion (LVO) ischemic stroke. At three months functional independence ranges between 33 and 60%, and mortality between 12 and 25% ^1–3^. Treatment effect size does not vary with age ^4^. However, few studies have reported outcomes beyond three-months and even fewer out to 5 years ^5–8^. These earlier studies identified factors that predicted mortality but their reported odds or hazard ratios are difficult to apply when making treatment decisions or conveying individual prognoses ^9^. Hazard ratios often vary over time, whilst using time specific hazard ratios introduces selection bias ^10^. Median survival cannot be used when mortality in a cohort of patients does not exceed 50%, and higher quantiles (e.g. 90^th^ centile) can be used but these take no account of subsequent events. Additionally, the difference between two survival curve quantiles, median or otherwise, does not represent extra life gained ^11^. For these reasons above adjusted survival curves, and their derived metrics, are preferred ^10^.

Restricted mean survival time (RMST) is defined as the area under a Kaplan-Meier curve or adjusted survival curve up to a specified cut-off time point. It has an advantage over hazard ratios and median survival in that it is a clinically relevant and stable metric that gives survival times up to the specified cutoff points ^9,12^. The probability of survival can be defined as the RMST divided by the time up to the said cut-off. RMST has the potential to provide clinicians, patients and families with realistic and interpretable expectations about recovery and survival following EVT.

We aimed to determine RMST at one, two and five years for anterior circulation LVO patients treated with EVT and quantify RMST differences within significant subgroups.

## Methods

This study was approved by the regional Health and Disability ethics committee (21/CEN/157) and a waiver of informed consent was granted. This study is reported following the Strengthening the Reporting of Observational Studies in Epidemiology guidelines checklist (Supplementary Material).

The study setting is a large comprehensive stroke center in New Zealand services 2.8 million people. Consecutive patients with anterior circulation LVO treated with EVT were identified from a mandatory national stroke therapy registry which has been active since May 2011. This registry contains data regarding patient demographics, medical co-morbidities, clinical features, treatment details and patient outcomes, as previously reported ^13^.

The attending clinical teams made all treatment decisions about eligibility for EVT. Baseline functional status was determined by the pre-stroke modified Rankin Scale score (mRS) ^14^. An Alberta Stroke program CT score (ASPECTS) of 0-5 identified those with large infarct cores, and 6-10 with smaller infarct cores ^15^. Stroke etiology was determined according to the Trial of Org 10 172 in Acute Stroke Treatment (TOAST) score ^16^. Reperfusion was graded using the modified Thrombolysis in Cerebral Infarction (mTICI) score ^17^. The primary outcome was survival at one, two and five years.

Separate from the national stroke therapy registry, data on all public and private health facility admissions were obtained from the National Minimum Data Set, a Ministry of Health supported database with demographic, process, and outcome data including international classification of diseases codes (ICD 9 and ICD 10) for all hospital episodes, and dates of death. Data transfer is completed within 3 weeks of hospital discharge with >99% of hospital episodes captured ^18^. Cases were excluded from analysis if they were missing key demographic or comorbidity data which might preclude analysis (Figure 1).

**Figure 1.**
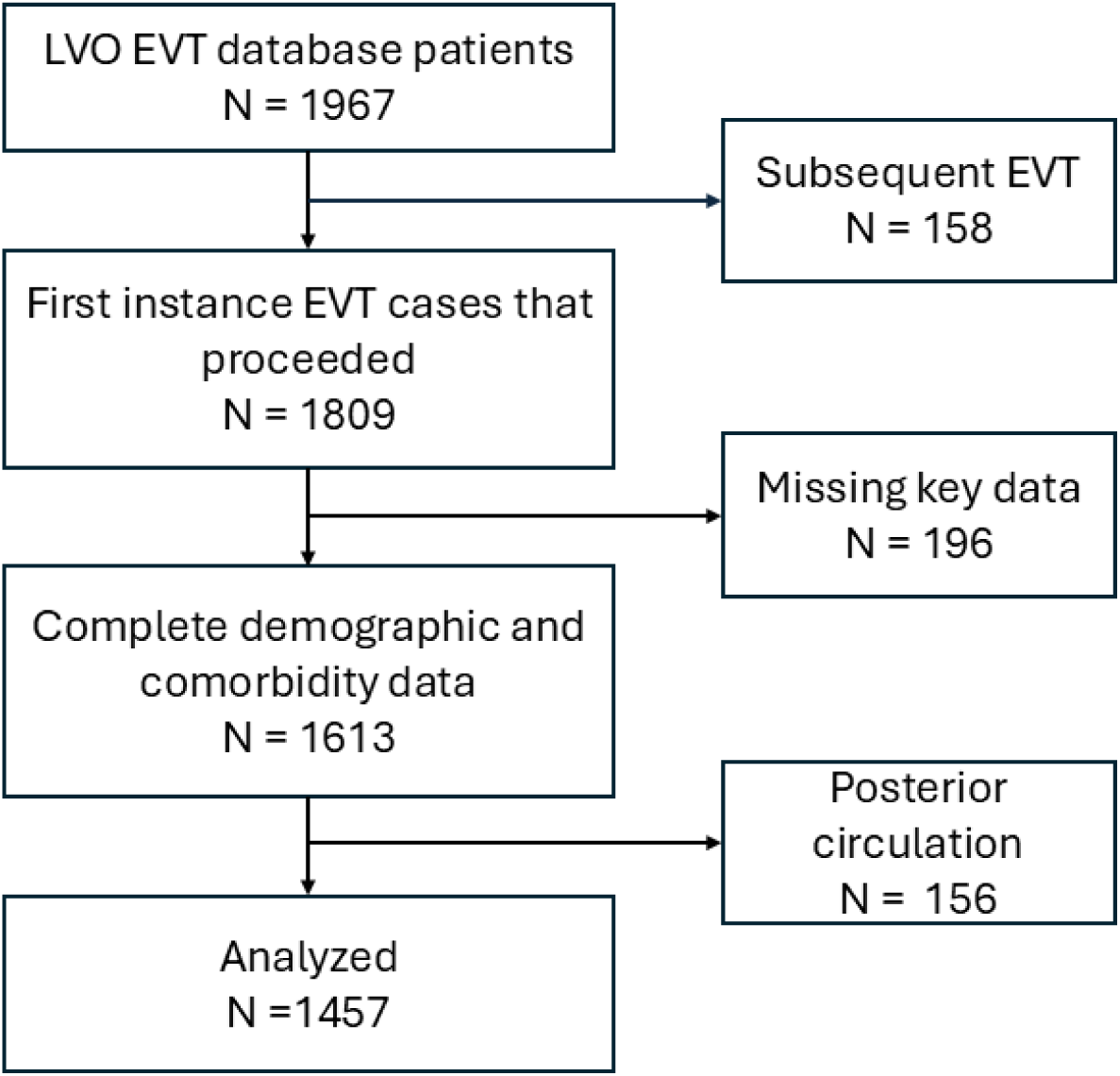
Cohort flow chart illustrating exclusion process for study patients. LVO – large vessel occlusion, EVT – endovascular thrombectomy.

International Classification of Diseases codes were extracted from the National Minimum Data Set data to classify co-morbidities and the Deyo-Charlson comorbidity index was calculated for each patient using the ‘comorbidity’ package for R ^19^. The Deyo-Charlson comorbidity index is a weighted score based on International Classification of Diseases coded co-morbidities that has been well validated and previously shown to be significant in predicting long-term post-EVT mortality ^6,20^.

## Statistics

Kaplan-Meier curves were generated from the data using the ‘survminer’ R package ^21^. For Cox regression analysis, all potential covariates were first assessed using univariate analysis, and those with p<0.1 were included for subsequent testing for proportionality of hazards as well as linearity for continuous variables prior to inclusion in the final Cox model. Stratification was used to overcome non-proportionality where necessary. The final Cox model was used to generate adjusted survival curves via the corrected group prognosis method (using the ‘survival’ and ‘adjustedCurves’ R packages) which were then used to find the RMST to compare subgroups ^22,23^. Unadjusted and adjusted RMST measures of accuracy were calculated using bootstrapping (n=1000). One, two and five years were selected as the cutoff time points for RMST.

For all analyses other than the univariate analysis, a p-value of less than 0.05 was considered statistically significant. All statistical analysis was performed in R (version 4.4.2), using R studio (version 2024.09.0+375, ‘Cranberry Hibiscus’) ^24,25^.

## Results

There were 1457 anterior circulation LVO patients treated with EVT (median age 71 [IQR: 60-80] years, 713 [49%] female) included in the analysis (Table 1). Median follow up was 8.9 years (95% CI 7.57 - inf) with maximum follow-up of 13 years. 1321 (90.1%) patients had been functionally independent (mRS 0-2) prior to the stroke. The median admission National Institutes of Health Stroke Scale Score (NIHSS) score was 16 (IQR: 11-20). 137 (9.3%) patients had large infarct cores on baseline imaging. 684 (47%) patients were treated with intravenous thrombolysis. The median time to reperfusion was 360min (IQR: 257-529) and 1296 (90%) patients achieved mTICI 2b-3. Death had occurred in 197 (14%) patients by three months, 270 (19%) by one year and 327 (22%) by two years and 415 (28%) by five years.

**Table 1:** Characteristics of anterior circulation EVT patients included for analysis.

| Characteristic | N = 1,457 |
| --- | --- |
| Age, med. (IQR) | 71 (60 - 80) |
| Female, n (%) | 713 (49%) |
| Pre-morbid mRS 0-2, n (%) | 1,320 (97%) |
| Missing | 98 |
| Admission NIHSS, med. (IQR) | 16 (11 - 20) |
| Missing | 17 |
| TOAST, n (%) |  |
| Cardioembolic | 767 (59%) |
| Large artery atherosclerosis | 252 (20%) |
| Other determined aetiology | 35 (2.7%) |
| Undetermined aetiology | 238 (18%) |
| Missing | 165 |
| Large core, n (%) | 137 (9.4%) |
| Thrombolysis, n (%) | 684 (47%) |
| Missing | 6 |
| Onset to needle, med. (IQR) | 148 (115 - 200) |
| Missing * | 784 |
| Time to recanalization, med. (IQR) | 360 (258 - 535) |
| Missing | 1 |
| Final TICl score, n (%) |  |
| 0 or 1 | 94 (6.5%) |
| 2a | 52 (3.6%) |
| 2b | 255 (18%) |
| 2c | 295 (20%) |
| 3 | 746 (52%) |
| Missing | 15 |
| Dead at 3 months, n (%) | 199 (14%) |
| Dead at 1 year, n (%) | 270 (19%) |
| Dead at 2 yrs, n (%) | 327 (22%) |
| Dead at 5 yrs, n (%) | 415 (28%) |
| mRS - modified Rankin Scale, NIHSS - National Institutes of Health stroke scale, TOAST - Trial of Org 10 172 in Acute Stroke Treatment, TICI - modified Thrombolysis in Cerebral Infarction score. |  |
| * Thrombolysis missing values include non-thrombolysed patients |  |

Kaplan-Meier analysis yielded median survival times of 7.6 years (95% CI 6.6-inf) for patients aged 70-79 years, 3.3 years (95% CI 2.8-4.6) for patients aged 80-89 years, and 0.6 years (95% CI 0.3-1.7) for patients aged 90-104 years. In each of the 10-year age brackets below 70 years, mortality did not exceed 50% by the censor date, precluding median analysis (Figure 2).

**Figure 2.**
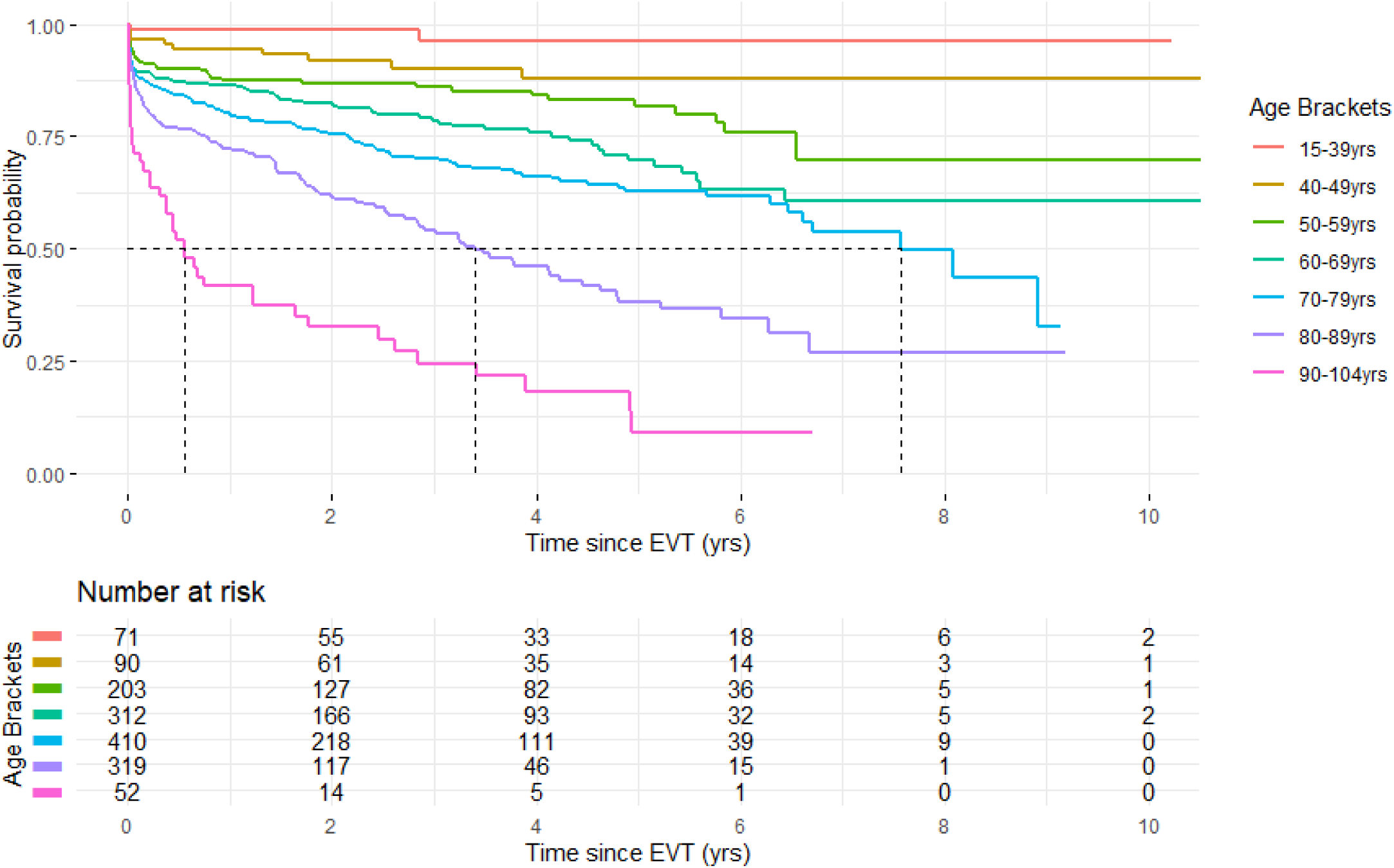
Kaplan-Meier curves for anterior circulation endovascular thrombectomy (EVT) patients by age brackets

After univariate analysis and tests for proportionality and linearity in the final Cox model covariates included were age, stroke hemisphere, infarct core size, admission NIHSS, ethnicity, Deyo-Charlson comorbidity index, thrombolysis, time to reperfusion, and final mTICI grade. The Cox model showed significant hazard ratios for death for patients aged 50 years or more (Table S1). Other significant co-variates were Deyo-Charlson index of comorbidities greater than two (HR 2.03 [95% CI 1.64-2.52], p <0.001), greater stroke severity at admission (NIHSS >15; HR 2.87 [95% CI 1.05-7.83], p =0.040), large infarct core at baseline (ASPECTS 0-5; HR 1.57 (95% CI 1.13-2.19), p =0.007), and little or no reperfusion (TICI score 0-2a; HR 1.9 (95% CI 1.43-2.53), p <0.001).

The unadjusted RMST for age brackets and associated probabilities for cut-off times are listed in Table 2. Patients aged 15-39 years had a one-year RMST of 0.98 years (95% CI 0.97-1.00), and a one-year survival probability of 98%. In contrast, those aged 90 years or older had one-year RMST of 0.52 years (95% CI 0.42-0.62) and a one-year survival probability of 52%. At the five-year cut-off, patients aged 15-39 years had a RMST of 4.86 years (CI 4.69 – 5.02), and a five-year survival probability of 97%, while those older aged 90 years or older had a RMST of 1.64 years (CI 1.17 – 2.11 years), and a five-year survival probability of 33%. The dynamic RMST curves showed marked leveling-off of survival for those aged over 80 years, and the curve for those aged 90 or older suggests a maximum RMST of 2 years over a longer period of follow up (Figure S1).

**Table 2:** Restricted mean survival time at one, two and five years after anterior circulation EVT by age brackets.

|  | Age Brackets |  |  |  |  |  |  |
| --- | --- | --- | --- | --- | --- | --- | --- |
|  | 15-39 yrs | 40-49 yrs | 50-59 yrs | 60-69 yrs | 70-79 yrs | 80-89 yrs | 90-104 yrs |
| One-year survival |  |  |  |  |  |  |  |
| RMST (years) | 0.98 | 0.93 | 0.89 | 0.88 | 0.83 | 0.74 | 0.52 |
| Probability (%) | 98 | 93 | 89 | 88 | 83 | 74 | 52 |
| SE | 0.01 | 0.02 | 0.02 | 0.01 | 0.01 | 0.02 | 0.05 |
| CI | 0.97-1 | 0.89-0.96 | 0.86-0.93 | 0.85-0.9 | 0.8-0.86 | 0.7-0.77 | 0.43-0.61 |
| Two-year survival |  |  |  |  |  |  |  |
| RMST (years) | 1.96 | 1.83 | 1.75 | 1.71 | 1.60 | 1.38 | 0.91 |
| Probability (%) | 98 | 91 | 87 | 85 | 80 | 69 | 45 |
| SE | 0.02 | 0.04 | 0.04 | 0.03 | 0.03 | 0.04 | 0.10 |
| CI | 1.92-2 | 1.74-1.91 | 1.67-1.82 | 1.64-1.77 | 1.54-1.66 | 1.31-1.45 | 0.71-1.1 |
| Five-year survival |  |  |  |  |  |  |  |
| RMST (years) | 4.86 | 4.39 | 4.10 | 3.96 | 3.60 | 2.93 | 1.64 |
| Probability (%) | 97 | 88 | 82 | 79 | 72 | 59 | 33 |
| SE | 0.08 | 0.15 | 0.12 | 0.10 | 0.09 | 0.11 | 0.24 |

| Age Brackets |  |  |  |  |  |  |  |
| --- | --- | --- | --- | --- | --- | --- | --- |
|  | 15-39 yrs | 40-49 yrs | 50-59 yrs | 60-69 yrs | 70-79 yrs | 80-89 yrs | 90-104 yrs |
| CI | 4.7-5.02 | 4.1-4.68 | 3.87-4.34 | 3.77-4.16 | 3.42-3.79 | 2.71-3.15 | 1.17-2.12 |

RMST - Restricted mean survival time, Probability - RMST/time point , SE - Standard error, CI - 95% Confidence interval

Adjusted RMST difference between patient subgroups with statistically significant hazard ratios are listed in Table 3. The RMST was determined from the adjusted survival curves shown in Figure 3. At the five-year cut-off patients with an independent baseline level of function had an absolute RMST gain of 0.77 years (CI 0.29-1.26 years, p = 0.002) compared with those with a poor baseline level of function. At the five-year cut-off patients with a lesser degree of comorbidity, as determined by a Deyo-Charlson Index of 0-2, had an absolute RMST gain of 0.71 years (CI 0.45-0.96 years, p < 0.001) compared with those with a greater degree of comorbidity. At the five-year cut-off, patients with large infarct cores on baseline imaging died six months earlier than those with smaller cores (0.48 years; 95% CI 0.10-0.87 years, p = 0.014). Patients with an admission NIHSS ≤15 also died six months earlier than those with higher admission NIHSS (0.49 years; CI 0.29-0.69 years, p < 0.001). Patients who achieved good final reperfusion (mTICI 2b-3) had an absolute RMST gain of 0.67 years (CI 0.35-1.00 years, p <0.001) at the five-year cut-off compared with those with poor reperfusion.

**Figure 3:**
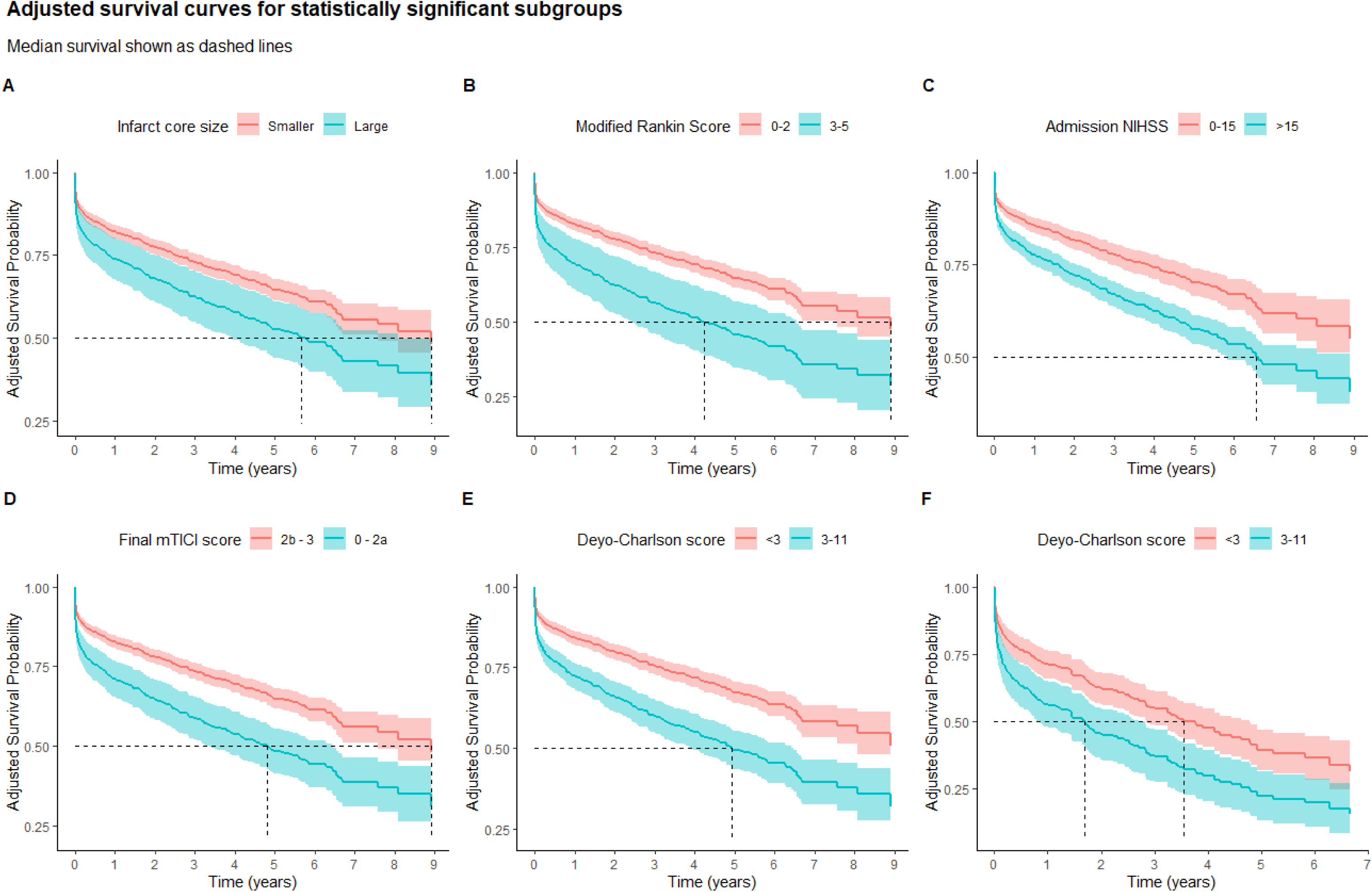
Adjusted survival curves for statistically significant subgroups used to determine the restricted mean survival time and median survival. Patients with large versus smaller infarct core size on baseline CT imaging (A), patients with baseline modified Rankin Score of 0-2 versus those with a score of 3-5 (B), patients with an admission NIHSS score of 0-15 versus those with a score greater than 15 (C), patients with a final modified TICI grade of 2b-3 versus those with a grade of 0-2a (D), patients with a low degree of comorbidity (Deyo-Charlson Index <3) on admission versus those with a higher degree comorbidity (Deyo-Charlson Index ≥3) (E), and patients aged over 80 with a low degree of comorbidity (Deyo-Charlson Index <3) versus those with a higher degree of comorbidity (Deyo-Charlson Index ≥3) (F). mTICI – modified treatment in cerebral infarction, NIHSS – National Institutes of Health Stroke Scale

**Table 3:** Difference in adjusted restricted mean survival between statistically significant subgroups.

|  | Absolute difference<br>(yrs) | CI | SE | p-value |
| --- | --- | --- | --- | --- |
| One-year cut-off |  |  |  |  |
| Baseline mRS 0-2 * | 0.11 | 0.03 - 0.19 | 0.04 | 0.005 |
| Deyo-Charlson Index 0-2 † | 0.10 | 0.06 - 0.14 | 0.02 | <0.001 |
| Admission NIHSS ≤15 ‡ | 0.07 | 0.04 - 0.09 | 0.02 | <0.001 |
| Smaller infarct core § | 0.07 | 0.01 - 0.13 | 0.03 | 0.025 |
| Final TICl 2b - 3 \|\| | 0.10 | 0.04 - 0.15 | 0.03 | <0.001 |
| Two-year cut-off |  |  |  |  |
| Baseline mRS 0-2 * | 0.25 | 0.08 - 0.42 | 0.09 | 0.004 |
| Deyo-Charlson Index 0-2 † | 0.23 | 0.14 - 0.31 | 0.04 | <0.001 |
| Admission NIHSS ≤15 ‡ | 0.15 | 0.09 - 0.22 | 0.03 | <0.001 |
| Smaller infarct core § | 0.16 | 0.02 - 0.29 | 0.07 | 0.02 |
| Final TICl 2b - 3 \|\| | 0.22 | 0.1 - 0.34 | 0.06 | <0.001 |
| Five-year cut-off |  |  |  |  |
| Baseline mRS 0-2 * | 0.77 | 0.28 - 1.27 | 0.25 | 0.002 |
| Deyo-Charlson Index 0-2 <sup>†</sup> | 0.71 | 0.45 - 0.96 | 0.13 | <0.001 |
| Admission NIHSS ≤15 <sup>‡</sup> | 0.49 | 0.29 - 0.69 | 0.10 | <0.001 |
| Smaller infarct core <sup>§</sup> | 0.48 | 0.1 - 0.87 | 0.20 | 0.014 |
| Final TICI 2b - 3 | 0.67 | 0.34 - 1.01 | 0.17 | <0.001 |

SE - Standard error, CI - Confidence interval, \* versus baseline mRS 3-5, † versus Deyo-Charlson index 3-11, ‡ versus Admission NIHSS >15, § versus large infarct core, || versus final mTICI of 0-2a

Adjusted median survival was only calculable for subgroups who achieved at least 50% adjusted mortality (Table S2). Upper confidence limits for lower risk groups were indeterminate. Adjusted median survivals for subgroups were as follows; patients with large infarct cores 5.67 years (CI 4.16-8.91 years); baseline mRS 3-5 4.23 years (CI 2.59-8.08 years); admission NIHSS greater than 15 6.55 years (CI 5.84-8.08 years); and final mTICI of 0-2a 4.80 years (CI 3.78 – 6.67 years). Adjusted survival curves show the subgroups where median survival could not be calculated (Figure 3).

Patients aged 80 and older, with a lesser degree of comorbidity (Deyo-Charlson score 0-2) had a five-year survival probability of 60% and an RMST of 3.0 years (CI 2.71 – 3.28 years) compared with those with a higher degree of comorbidity (survival probability of 43.8%, RMST 2.19 years [CI 1.74 – 2.64 years]). The five-year RMST difference between the two comorbidity levels was 0.81 years (CI 0.27 – 1.33 years; p = 0.003). The median survival for patients aged 80 and older with a lesser degree of comorbidity was 3.55 years (CI 2.83 – 4.81 years) compared with those with a higher degree of comorbidity (1.69 years [CI 0.71 – 2.71 years]). Patients aged 80 years and older with an admission NIHSS of ≤ 15 had a five-year survival probability of 65.0% with an adjusted RMST of 3.25 years (CI 2.92 – 3.58 years) compared with those with an admission NIHSS > 15 (survival probability 47.4%, adjusted RMST 2.37 years [CI 2.06 – 2.68 years]). The five-year RMST difference between these NIHSS groups was 0.88 years (CI 0.43 – 1.33 years; p <0.001). For patients aged 80 and older with an admission NIHSS of ≤ 15 the median survival was 4.63 years (CI 3.24 – 6.28 years), compared with admission NIHSS of > 15 (median survival 1.88 years [CI 1.47 – 2.84 years]).

## Discussion

This study has reported clinically useful estimates of survival time at one, two and five years in a large cohort of LVO patients treated with EVT. The novel aspect of this study was the use of RMST, which provides a quantifiable and comparable metric to assess survival across different ages and between subgroups. RMST is easy to communicate, captures survival nuances, and can provide absolute differences that are not possible when outcomes are measured using median survival alone. In each of the 10-year cohorts less than 70 years of age, fewer than half of the patients died, meaning a median survival could not be calculated. In these groups, RMST provided both an average survival and the probability of surviving up to a selected cut-off time point, which is more intuitive in those with lower mortality rates. In patients aged 80-89 years, median survival and RMST are complementary, with median survival of 3.3 years and five-year RMST of 2.93 years. In patients aged 90 or older, median survival was 0.6 years while the five-year RMST was 1.64 years (95% CI 1.17-2.11). This reflects median survival times being weighted by early deaths and not capturing the later slowing mortality rate seen in the Kaplan-Meier curve.

The use of RMST in survival studies to date has been limited, and to our knowledge there have been no stroke studies utilizing this method ^26,27^. Some *post-hoc* analyses have applied RMST to previously published work ^28^. The challenge of using RMST in prospective studies is selecting appropriate cutoff time points. A key advantage in the application of RMST to retrospective studies is that RMST differences between comparator groups (e.g. age cohorts or those with and without large infarct cores) have operating characteristics akin to the log-rank test and can quantify differences where there are non-proportional hazards ^9,10^.

Sub-group analysis using adjusted RMST quantified the absolute survival differences for groups with significant hazard ratios such as baseline level of function, comorbidity, admission NIHSS, baseline core size, and find reperfusion grade. Survival differences for the various subgroups ranged from six to nine months, meaning decisions based on the likelihood of long-term survival should probably not be based on one factor alone. For all subgroups the adjusted survival curves diverged although this was most pronounced for baseline level of function, comorbidity, and final recanalization. Thus, when considering an LVO patient for EVT with multiple risk factors, those with poor baseline function and higher comorbidity can be expected to have a significantly shorter survival especially if final reperfusion is poor.

Anterior circulation LVO patients with large infarct cores on baseline imaging had a reduction in five-year adjusted RMST of almost six months, compared with those with smaller infarct cores. This mortality excess is seen at one year and extends out to five years. It is surprising to find that this did not have as big an impact on long-term survival as some other factors, but this should be taken in context of the study limitations. The similarity in survival gains between large and smaller core size, and low and higher admission NIHSS likely reflects the collinearity between these metrics.

For patients aged 80 years and older, those with a lower degree of comorbidity lived nearly 10 months longer on average than those with a higher degree of comorbidity. Whilst the median survival by degree of comorbidity for those aged 80 years or older also reflects this survival difference, on their own the wide confidence intervals make clinical application more challenging. Similarly, admission NIHSS has a significant effect on survival in those aged 80 years and older, with those with a NIHSS ≤ 15 having a 10.5-month survival advantage over those with an admission NIHSS > 15. Median survival confirmed the relationship but exaggerated survival gain or loss.

This study has limitations. This is an observational study in a highly selected population of LVO patients treated with EVT within a single center which limits the generalizability of these findings. The effect of individual comorbidities on survival were not assessed in this study. Posterior circulation LVO patients were not included as they are a clinically distinct group with a different survival profile.

Strengths include the large number of patients and the application of a novel survival measure. For the Cox regression we used the Deyo-Charlson comorbidity index due to its prior validation, extensive use, and easy derivation from ICD codes. Use of a comorbidity index takes into account the multiplicative effect of comorbidities and avoids multi-collinearity.

We have used RMST to estimate a clinically useful estimate of survival of anterior circulation LVO patients treated with EVT. RMST indicates that survival of patients of advanced age is likely longer than median survival suggests. Furthermore, we have quantified the survival differences for various important subgroups. RMST conveys likely prognosis accurately and in a way that can be easily understood by patients, families, and clinicians. This is especially important when considering EVT in patients of advanced age. RMST should be considered as a useful metric in future studies examining survival, especially when difference in survival is expected to be close, or rates of death are low.

## Data Availability

The data used for this study is from several different national registries and databases. Ethics approval for use of this health data was contingent on us not sharing this raw data with anyone other than those stipulated on the ethics application.

## Acknowledgements

There are no acknowledgements

## Supplemental Material

Tables S1–S2

Figure S1

Strengthening the Reporting of Observational Studies in Epidemiology guidelines checklist

## Non-standard Abbreviations and Acronyms

ASPECTS: Alberta Stroke Program Early Computer Tomography Score
EVT: endovascular thrombectomy
LVO: large vessel occlusion
mRS: modified Rankin score
mTICI: modified treatment in cerebral infarction
NIHSS: National Institutes of Health Stroke Scale
RMST: Restricted Mean Survival Time

